# Association of daily steps with incident non-alcoholic fatty liver disease: Evidence from the UK Biobank cohort

**DOI:** 10.1101/2024.05.17.24307527

**Authors:** Evelynne S. Fulda, Laura Portas, Charlie Harper, David Preiss, Derrick Bennett, Aiden Doherty

## Abstract

**Background:** Low physical activity (PA) has been shown to be associated with higher risk of non-alcoholic fatty liver disease (NAFLD). However, the strength and shape of this association are currently uncertain due to a reliance on self-reported PA measures. This report aims to investigate the relationship of PA with NAFLD using accelerometer-derived step count from a large prospective cohort study.

**Methods:** The wrist-worn accelerometer sub-study of the UK Biobank (N=∼100,000) was used to characterise median daily step count over a seven-day period. NAFLD cases were ascertained via record linkage with hospital inpatient data and death registers or by using a measure of liver fat from imaging. Cox proportional hazards models were employed to assess the association between step count and NAFLD, adjusting for age, sociodemographic, and lifestyle factors. Mediation analyses were conducted.

**Results:** Among 91,031 participants (709,440 person-years of follow-up), there were 762 incident NAFLD cases. Higher step count was log-linearly and inversely associated with risk of NAFLD. A 1000 step increase (representing 10 minutes of walking) was associated with a 12% (95% CI: 10%–14%) lower hazard of NAFLD. When using imaging to identify NAFLD, a 1,000-step increase was associated with a 6% (95% CI: 6%–7%) lower risk. There was evidence for some mediation by adiposity.

**Conclusion:** Physical activity, a modifiable risk factor, is log-linearly and inversely associated with NAFLD. This association was only partially explained by adiposity. These findings from a large cohort study may have important implications for strategies to lower NAFLD risk.

## INTRODUCTION

Non-alcoholic fatty liver disease (NAFLD) is the most common cause of chronic liver disease with an estimated global prevalence of 30%.^1^ It is associated with multiple comorbidities, is a leading cause of cirrhosis and liver failure, and is predicted to be the most common indication necessitating liver transplantation over the next decade.^2^ Importantly, NAFLD prevalence is increasing, primarily as a result of population-wide lifestyle changes.^3^ Thus, lifestyle modification, such as changes in physical activity, may represent an opportunity for primary prevention of NAFLD. For example, prospective studies have suggested that higher levels of physical activity are associated with a lower risk of incident NAFLD,^4–12^ with clinical trials demonstrating evidence for resolution of fatty liver with increased exercise.^13^

However, the current physical activity and NAFLD evidence base has several limitations. First, most prior studies used subjective measures of activity. These measures are prone to substantial measurement error and bias, do not measure activity continuously, are difficult to compare between subjects, and only capture intentional physical activity.^14^ In relation to this, physical activity may be characterised in unintuitive ways or as relative measures of activity, which are population-dependent. Finally, there is a limited understanding of how adiposity, hypertension, medication use, or diabetes potentially mediate the association between physical activity and NAFLD risk. Prior studies have tended to treat these covariates as only confounders, without considering their intermediary role.^15^

Using comprehensive data from the UK Biobank, we aimed to 1) investigate the association between accelerometer-measured physical activity characterised as median daily step count and incident NAFLD and 2) consider how potential factors mediate this association.

## METHODS

### Study design and participants

The UK Biobank is a prospective cohort study that enroled approximately 500,000 adults between 2006 and 2010 across England, Scotland, and Wales.^16^ Full information on the UK Biobank study design, recruitment strategy, and data collection protocols have been previously described.^16^ Briefly, participants were eligible to take part in the study if they were aged 40-69 and resided within driving distance of an assessment centre at the time of recruitment. All participants provided written informed consent and completed a baseline visit involving collection of sociodemographic, lifestyle, and health information. Excepting cases of withdrawal of consent, migration, and/or death, participants have been passively followed continuously since enrolment.

To derive the main analysis cohort, participants without accelerometer data or with accelerometer data not meeting quality standards, prevalent liver disease (NAFLD and other liver diseases assessed using record linkage; International Classification of Diseases [ICD] codes outlined in supplemental table S1) or history of alcohol misuse (assessed using record linkage; ICD codes outlined in supplemental table S1) at the time of accelerometer wear, or missing covariate data were excluded.

### Physical activity assessment

A random selection of participants with a valid email address (N=236,519) were invited to wear a wrist-worn accelerometer for seven days.^17^ 45% of participants volunteered to take part in this sub-study, with data collection commencing in June 2013 and concluding in December 2015.^17^ During this period, participants were mailed an Axivity AX3 triaxial accelerometer. The accelerometer was programmed to automatically capture data two days after postage at 10:00 am and to measure continuously for a 7-day period. Participants were instructed to wear the accelerometer without interruption on their dominant wrist and to conduct their normal activities.

From the raw accelerometer data, daily step count was quantified using a hybrid self-supervised machine learning and peak detection algorithm.^18^ This algorithm was validated against reference video measurements from personal wearable cameras.^18^ The median daily step count over the seven-day wear period was obtained. Step count was categorised into quartiles to look at the overall shape of the association. As the shape was log-linear, continuous step count was also assessed. A cadence (walking pace) of 100 steps per minute is considered moderate^19^; thus, the continuous step count variable is presented as per 1,000-step increase, representing on average ten more minutes of walking per day.

Physical activity was additionally assessed among all participants using the International Physical Activity Questionnaire (IPAQ). This questionnaire assessed how many days per week a participant completed more than ten minutes each of walking, moderate-level activity, or vigorous-level activity, as well as how many minutes per day a participant typically spends on each activity type based on a participant’s self-report. These responses were then used to derive total Metabolic Equivalent of Task (MET)-minutes per week for all activity according to IPAQ guidelines,^20^ which was then grouped by quartiles.

### NAFLD ascertainment

NAFLD cases were identified through linkage to routinely collected hospital admission and national death registry data using ICD-10 (K75.8 and K76.0) and ICD-9 (571.5, 571.8, and 571.9) codes. This analysis linked to both primary and secondary ICD codes to capture participants admitted both for and with NAFLD and to capture NAFLD as either an underlying or contributory cause of death.

A subset of UK Biobank participants underwent abdominal magnetic resonance imaging (MRI), with subsequent processing of imaging-derived phenotypes. To assess how the NAFLD ascertainment method may affect findings, among participants from the main analysis with both accelerometer and imaging data, NAFLD was defined using proton density fat fraction (PDFF) derived from MRI of the liver. A PDFF >5.5% is considered to be indicative of NAFLD.^21^ Imaging of UK Biobank participants commenced on 30 April 2014 and is still underway at the time of the present analysis (March 2024).

Participants were censored at the time of their first NAFLD diagnosis, death, withdrawal from the study, or end of follow-up. Record linkage censoring dates varied by region: 31 October 2022 for participants in England, 31 August 2022 for participants in Scotland, and 31 May 2022 for participants in Wales.

### Covariates

At the baseline visit, information relating to multiple covariates was collected based on self-report (unless otherwise specified). Variables were categorised subsequently as follows: sex assigned at birth (male, female); ethnicity (white, non-white); Townsend deprivation index (UK population quintile; from postcode); educational attainment (school leaver, further education, higher education); alcohol consumption (≥3 times/week, <3 times/week, never); smoking status (current/former, never); fruit and vegetable intake (<3, 3-4.9, 5-7.9, ≥8 servings/day); self-rated health (poor, fair, good, excellent); body mass index ([BMI] continuous); systolic blood pressure (continuous; from automatic or manual sphygmomanometer); use of cholesterol-lowering medication, blood pressure medication, and/or insulin (on medication, not on medication); number of unique hospital episodes (continuous); and health conditions based on algorithmic-definitions in the UK Biobank (cancer) or ICD codes outlined in supplemental table S1 (diabetes, hypertension, or chronic lower respiratory disease). For covariates that allowed participants to respond with “I don’t know” or “Prefer not to answer”, these responses were interpreted as missing.

### Statistical Analysis

Cox proportional hazards regression models using age at the timescale were employed to assess the association between device-measured step count and risk of incident NAFLD. The model was adjusted for sex, ethnicity, Townsend deprivation index, educational attainment, alcohol consumption, smoking status, and fruit/vegetable consumption (the adjusted model). Hazard ratios are presented with their accompanying 95% confidence intervals (CI) using floating absolute risks where appropriate to present group-specific risks and their variances.^22^ Analyses were repeated using quartiles of MET-minutes per week as the exposure.

A mediation analysis was conducted for covariates thought to lie on the mechanistic pathway between step count and NAFLD. The included variables were BMI, hypertension, medication use, and diabetes. The effect of mediation was assessed in three ways: reduction in Χ^2^, calculation of risk mediated, and causal mediation. To investigate the reduction in Χ^2^, a likelihood ratio test was performed by fitting the adjusted model with and without step count (Model A). Next, a likelihood ratio test was performed by fitting the adjusted model with the addition of the mediator(s) with and without step count (Model B). Finally, the reduction in the likelihood ratio Χ^2^ test statistic from Model A to Model B was assessed. This was followed by an estimation of percent of risk mediated (PRM), as adapted from the Global Burden of Metabolic Risk Factors for Chronic Diseases Collaboration^15^:

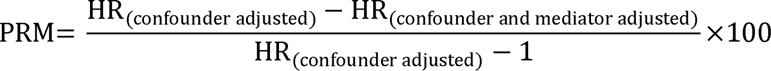

Each mediator was assessed individually, in all combinations of two, and all four together.^15^ Finally, a formal causal mediation analysis was conducted for each mediator individually using the *mediate* package in Stata. The total effect (the difference in NAFLD risk if everyone exercised like those in the most active quarter compared to the least) was quantified and split into the natural direct effect (the effect of the exposure on the outcome independent of the mediator) and the natural indirect effect (the effect of the exposure on the outcome due to the mediator). We report the proportion mediated, which is calculated as:

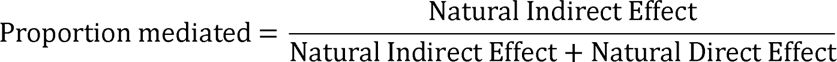

This method involves a counterfactual framework, as described elsewhere.^23^

Four sensitivity analyses were conducted. The primary analysis relies on linkage with hospital and death records; therefore, the observed association may be due to physical activity being associated with risk of hospitalization or death. To account for this potential ascertainment bias, the first sensitivity analysis excluded participants with self-reported poor health or with potentially relevant diseases (cancer, chronic lower respiratory disease, diabetes, and hypertension) and additionally adjusted for the unique number of hospital episodes. The second sensitivity analysis excluded participants with events in the first five years of follow-up to consider the potential for reverse causation. For the third sensitivity analysis, models from the main analysis were repeated using either PDFF-defined NAFLD or record linkage-defined NAFLD among participants with both accelerometer and imaging data. Participants with prevalent NAFLD based on PDFF were additionally excluded from this analysis. As imaging visits were conducted after accelerometer wear for most participants, it was not possible to exclude all prevalent PDFF-defined NAFLD cases. Therefore, to allow for a cross-sectional analysis, the relationship between step count and log-transformed PDFF (transformed due to skew of distribution) was assessed using multivariable linear regression. The final sensitivity analysis sought to investigate the potential for unmeasured and residual confounding by calculating an E-value for the main analysis. The E-value quantifies the necessary strength of association for an unmeasured or inadequately measured confounder to explain away the association between the exposure and outcome, after adjustment for measured confounders.^24^

Normally distributed continuous variables are presented as mean (standard deviation [SD]), non-normally distributed continuous variables are presented as median (interquartile range [IQR]), and categorical variables are presented as n (%). A two-sided alpha level of 0.05 was considered statistically significant.

Data were extracted from the UK Biobank Research Analysis platform using pyspark, with methods as previously described.^17^ Data were pre-processed using R, version 4.3.1 (R Core Team, Austria). All statistical analyses were conducted using Stata, version 18.0 (StataCorp, USA). Results are reported according to the STROBE guidelines (supplemental table S2).

## RESULTS

### Baseline Characteristics

After excluding 12,328 participants, 91,031 participants were included in the present analysis (exclusion flow diagram in supplemental figure S1). The median age of participants was 63.4 (IQR: 56.2–68.5) years, 88,320 (97%) participants were white, and 51,647 (57%) participants were female (Table 1). Compared to participants in the first (lowest) step count quarter, participants in the fourth (highest) step count quarter were more likely to be younger (62.7 [IQR: 56.0–67.8] years vs. 64.0 [IQR: 56.6–69.3] years), to consume alcohol more frequently (54% drank ≥3 times/week compared to 42%), and to have lower BMI (25.6 [SD: 3.7] kg/m^2^ vs. 28.2 [SD: 5.4] kg/m^2^).

**Table 1.** Baseline participant characteristics, overall and by median daily step count quarter.

|  | 0-6,999 steps<br>N=22,764 | 7,000-9,269 steps<br>N=22,753 | 9,270-11,891 steps<br>N=22,760 | 11,892-41,189 steps<br>N=22,754 | 0-41,189 steps<br>N=91,031 |
| --- | --- | --- | --- | --- | --- |
| **Median daily step count** | 5259 (1393) | 8150 (647) | 10490 (749) | 14783 (2763) | 9670 (3845) |
| **MET-min/week** | 1333 (570-2652) | 1635 (796-3066) | 1857 (946-3416) | 2319 (1235-4158) | 1773.0 (857-3339) |
| **Age at entry, years** | 64.0 (56.6-69.3) | 63.3 (56.0-68.6) | 63.4 (56.2-68.4) | 62.7 (56.0-67.8) | 63.4 (56.2-68.5) |
| **Female sex** | 13,067 (57.4%) | 13,129 (57.7%) | 12,934 (56.8%) | 12,517 (55.0%) | 51,647 (56.7%) |
| **White ethnicity** | 22,011 (96.7%) | 22,058 (96.9%) | 22,124 (97.2%) | 22,127 (97.2%) | 88,320 (97.0%) |
| **Townsend deprivation index quintile** |  |  |  |  |  |
| Least deprived | 11,132 (48.9%) | 11,992 (52.7%) | 11,726 (51.5%) | 11,410 (50.1%) | 46,260 (50.8%) |
| Second Quarter | 6,623 (29.1%) | 6,447 (28.3%) | 6,614 (29.1%) | 6,531 (28.7%) | 26,215 (28.8%) |
| Third Quarter | 1,782 (7.8%) | 1,653 (7.3%) | 1,687 (7.4%) | 1,857 (8.2%) | 6,979 (7.7%) |
| Fourth Quarter | 2,362 (10.4%) | 2,024 (8.9%) | 2,047 (9.0%) | 2,224 (9.8%) | 8,657 (9.5%) |
| Most deprived | 865 (3.8%) | 637 (2.8%) | 686 (3.0%) | 732 (3.2%) | 2,920 (3.2%) |
| **Highest education** |  |  |  |  |  |
| School leaver | 11,129 (48.9%) | 9,918 (43.6%) | 9,159 (40.2%) | 8,763 (38.5%) | 38,969 (42.8%) |
| Further education | 3,139 (13.8%) | 3,079 (13.5%) | 2,897 (12.7%) | 2,980 (13.1%) | 12,095 (13.3%) |
| Higher education | 8,496 (37.3%) | 9,756 (42.9%) | 10,704 (47.0%) | 11,011 (48.4%) | 39,967 (43.9%) |
| **Alcohol consumption** |  |  |  |  |  |
| 3+ times/week | 9,475 (41.6%) | 11,003 (48.4%) | 11,791 (51.8%) | 12,289 (54.0%) | 44,558 (48.9%) |
| <3 times/week | 11,673 (51.3%) | 10,550 (46.4%) | 9,848 (43.3%) | 9,369 (41.2%) | 41,440 (45.5%) |
| Never | 1,616 (7.1%) | 1,200 (5.3%) | 1,121 (4.9%) | 1,096 (4.8%) | 5,033 (5.5%) |
| **Never smoker** | 12,400 (54.5%) | 13,223 (58.1%) | 13,453 (59.1%) | 13,435 (59.0%) | 52,511 (57.7%) |
| **Fruit and vegetable consumption** |  |  |  |  |  |
| <3 servings/day | 1,114 (4.9%) | 727 (3.2%) | 642 (2.8%) | 625 (2.7%) | 3,108 (3.4%) |
| 3-4.9 servings/day | 3,328 (14.6%) | 3,000 (13.2%) | 2,774 (12.2%) | 2,506 (11.0%) | 11,608 (12.8%) |
| 5-7.9 servings/day | 8,322 (36.6%) | 8,440 (37.1%) | 8,352 (36.7%) | 8,183 (36.0%) | 33,297 (36.6%) |
| >8 servings/day | 10,000 (43.9%) | 10,586 (46.5%) | 10,992 (48.3%) | 11,440 (50.3%) | 43,018 (47.3%) |
| **Self-reported health** |  |  |  |  |  |
| Poor | 1,137 (5.0%) | 451 (2.0%) | 321 (1.4%) | 231 (1.0%) | 2,140 (2.4%) |
| Fair | 4,841 (21.3%) | 3,391 (14.9%) | 2,976 (13.1%) | 2,623 (11.5%) | 13,831 (15.2%) |
| Good | 13,061 (57.4%) | 13,853 (60.9%) | 13,955 (61.3%) | 13,982 (61.4%) | 54,851 (60.3%) |
| Excellent | 3,725 (16.4%) | 5,058 (22.2%) | 5,508 (24.2%) | 5,918 (26.0%) | 20,209 (22.2%) |
| **Body mass index, kg/m<sup>2</sup>** | 28.2 (5.4) | 26.7 (4.3) | 26.1 (4.0) | 25.6 (3.7) | 26.7 (4.5) |
| **Medication use†** | 6,629 (29.1%) | 5,094 (22.4%) | 4,741 (20.8%) | 4,003 (17.6%) | 20,467 (22.5%) |
| **Diabetes** | 1,165 (5.1%) | 601 (2.6%) | 436 (1.9%) | 343 (1.5%) | 2,545 (2.8%) |
| **Hypertension** | 4,212 (18.5%) | 2,929 (12.9%) | 2,546 (11.2%) | 2,115 (9.3%) | 11,802 (13.0%) |

Data are presented as mean (standard deviation) for normally distributed continuous measures, as median (interquartile range) for non-normally distributed continuous measures, and n (%) for categorical measures
†Use of cholesterol-lowering medication, blood pressure medication, and/or insulin

### Associations between daily step count and incident NAFLD

Over a median follow-up time of 7.9 (IQR: 7.3–8.4) years (709,440 person-years), there were 762 incident NAFLD cases identified using record linkage. Only 43 participants were admitted with NAFLD as the primary hospital diagnosis. The next most common primary diagnoses were R10.1 (pain localised to upper abdomen; n=21) and K80.1 (calculus of gallbladder with other cholecystitis; n=21). Further, only one participant had NAFLD listed as their primary death cause. After adjustment for confounders, compared to participants in the first step count quarter, participants in the fourth step count quarter had a hazard ratio of 0.35 (95% CI: 0.29–0.43; likelihood ratio test for trend Χ^2^=129, p<0.001; Figure 1). A 1,000-step higher step count (10 minutes of walking) was associated with a 12% lower hazard of incident NAFLD (0.88, 0.86–0.90). Sequential adjustment of confounders is shown in supplemental figure S2. When using self-report to characterise physical activity, participants in the fourth MET-min/week quarter had a 39% lower hazard of NAFLD than those in the first (0.61, 0.51–0.72; supplemental figure S3).

**Figure 1.**
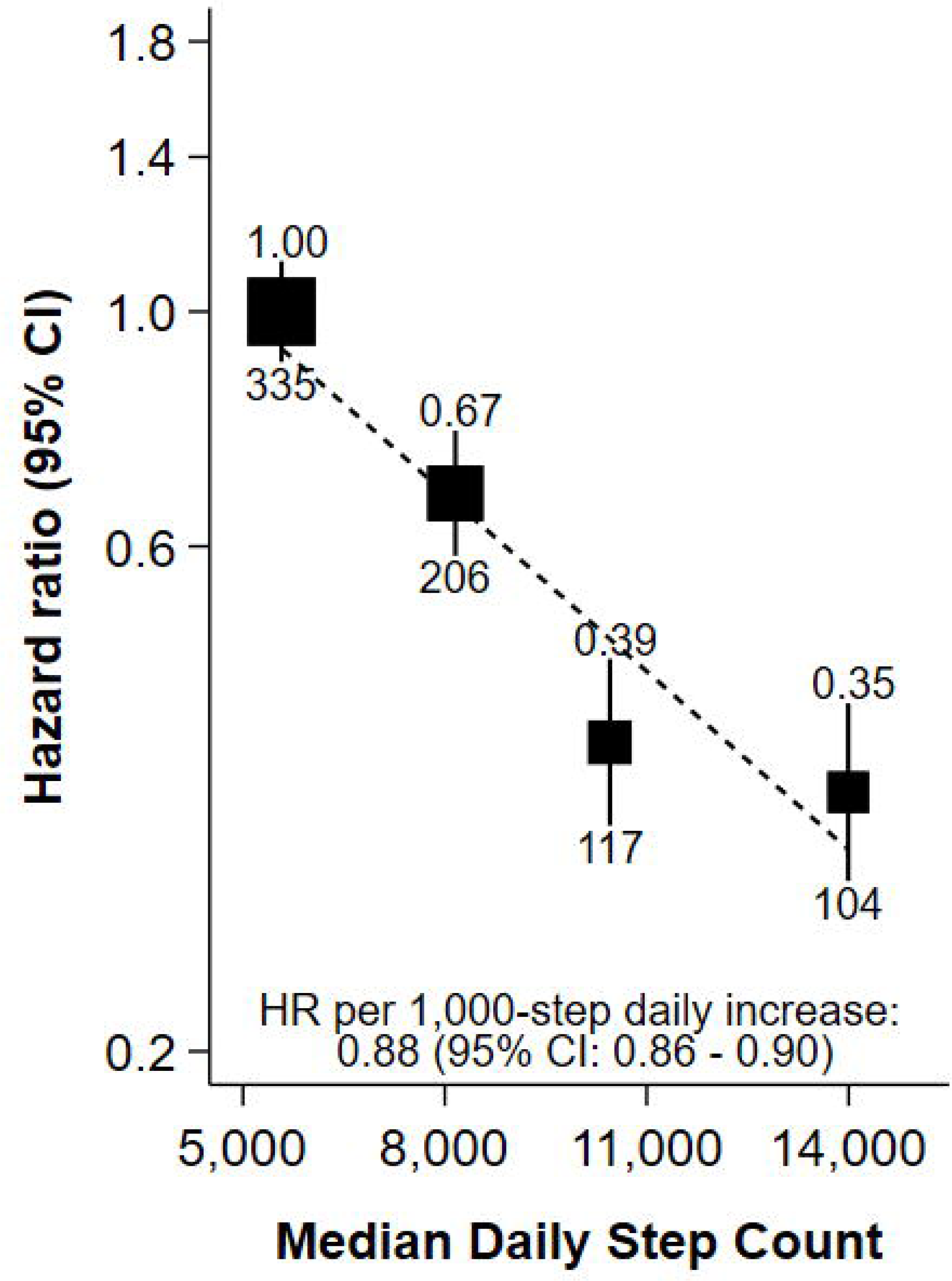
Association of quartiles of accelerometer-measured daily step count with risk of NAFLD after a median of 7.9 years of follow-up in 91,031 UK Biobank participants. Adjusted for sex, ethnicity, Townsend deprivation index, educational attainment, alcohol consumption, smoking status, fruit and vegetable consumption and using age as the time scale; the number above each vertical line is the HR, and the number below each vertical line is the number of events.

### Mediation analysis

Adding BMI into the model noticeably attenuated the association between step count and risk of NAFLD (0.93, 0.91–0.95), with a reduction in the likelihood ratio test statistic of 66% and a percent risk mediated (PRM) of 39% (Figure 2). Adding hypertension, medication use, or diabetes into the model also attenuated the association, though with a smaller effect (hypertension: 0.89, 0.87–0.91, reduction in Χ^2^: 18%, PRM: 9%; medication use: 0.89, 0.87–0.91, reduction in Χ^2^: 17%, PRM: 8%; diabetes: 0.89, 0.87–0.91, reduction in Χ^2^: 16%, PRM: 8%; Figure 2). In models including multiple mediators, the attenuation of the hazard ratio, reduction in Χ^2^, and PRM was largely driven by the inclusion of BMI. Further, in causal mediation models, BMI showed the highest proportion of mediation (29%; Figure 2). When restricting analyses to only include participants with obesity (BMI≥30 kg/m^2^), the association between step count and hazard of NAFLD was attenuated (0.92, 0.89–0.95) and no longer showed a clear log-linear dose-response pattern (supplemental figure S4).

**Figure 2.**
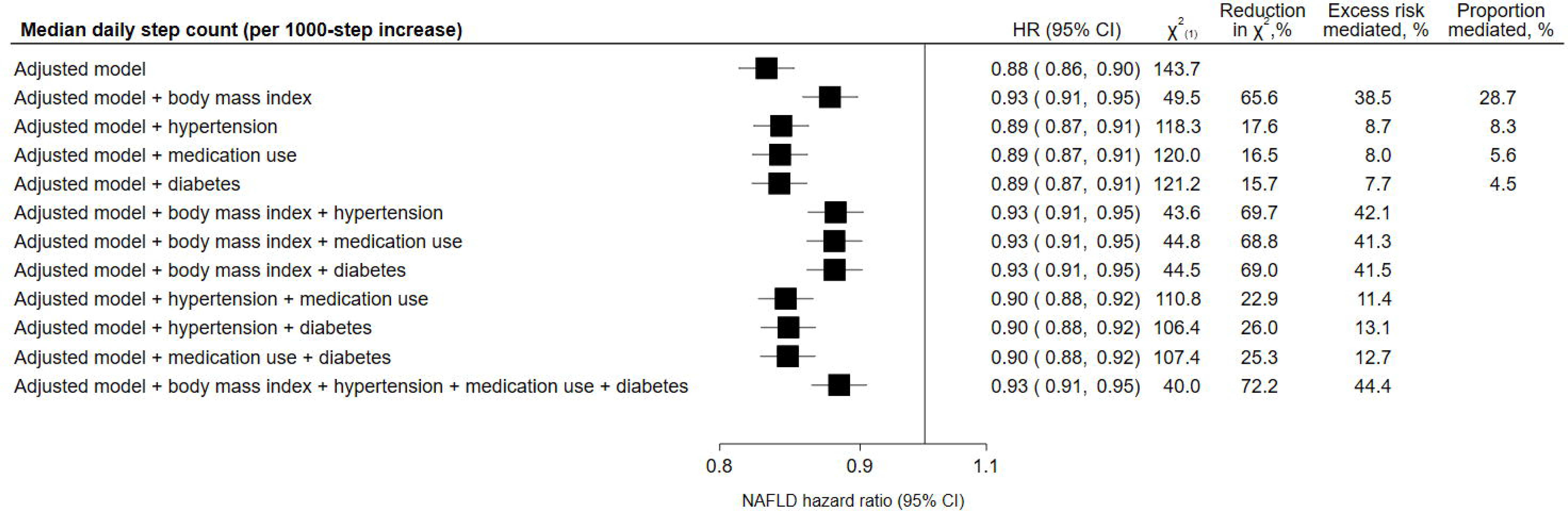
Association of accelerometer-measured daily step count with risk of NAFLD, inclusion of potential mediators. Adjusted model: controlled for sex, ethnicity, alcohol consumption, smoking status, educational attainment, Townsend deprivation index, fruit and vegetable consumption and using age as the time scale Reduction in Χ^2^ (in relation to adjusted model), quantifies the amount the observed association is due to the mediator(s) Risk mediated, quantifies the percent change in the effect estimate after addition of the mediator(s) Proportion mediated, quantifies the proportion of the total effect that is due to the mediator.

### Sensitivity analyses

Exclusion of events that occurred during the first five years of follow-up (389 events) or exclusion of individuals with ill health and controlling for number of hospitalizations (n=26,580) both marginally attenuated the association between step count and hazard of incident NAFLD: HR per 1,000-step increase (95% CI): 0.90 (0.87–0.92) and 0.90 (0.88–0.93), respectively (shape plots shown in supplemental figure S5).

Among participants with accelerometer and imaging data (N=15,689), there were 3,276 NAFLD cases based on PDFF and 107 NAFLD cases based on record linkage, with median follow-up times of 7.6 (IQR: 6.9–8.1) and 7.8 (IQR: 7.4–8.3) years, respectively. The association between step count and risk of NAFLD was attenuated compared to the main analysis for both PDFF-ascertained NAFLD (0.94, 0.93–0.94) and record linkage-ascertained NAFLD (0.94, 0.89–0.99). Figure 3 investigates the shape of these associations by step count quartile. Step count was inversely associated with PDFF in linear regression analyses: each 1,000-step increase in step count was associated with a 2% lower PDFF value on average. The association between step count quartile and log(PDFF) is shown in supplemental figure S6.

**Figure 3.**
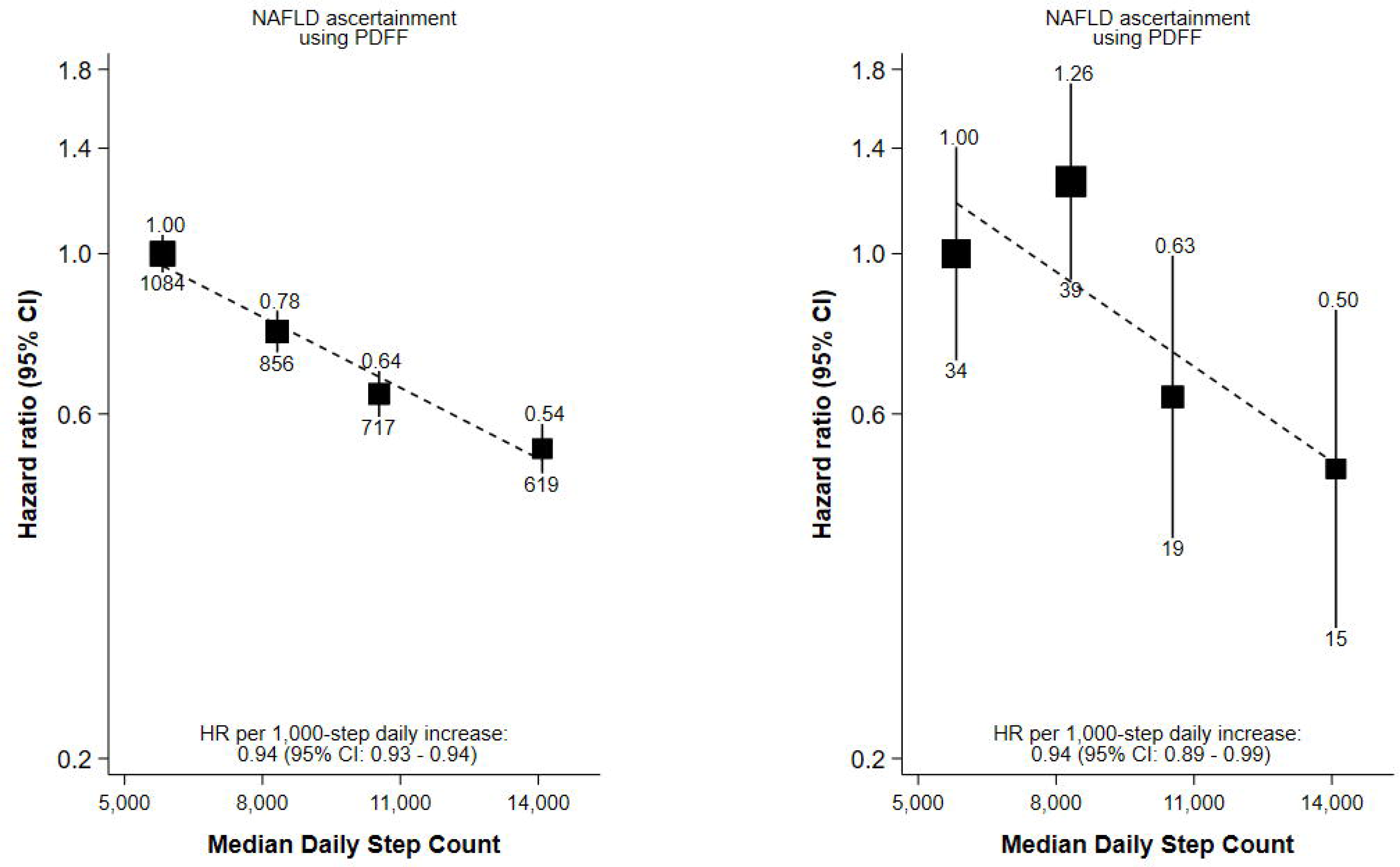
Association of quartiles of accelerometer-measured daily step count with risk of NAFLD in 15,689 UK Biobank participants with both accelerometer and imaging data. Adjusted for sex, ethnicity, Townsend deprivation index, educational attainment, alcohol consumption, smoking status, fruit and vegetable consumption and using age as the time scale; the number above each vertical line is the HR, and the number below each vertical line is the number of events.

The E-value for the main analysis hazard ratio of 0.88 (0.86–0.90) was 1.52 with an interval bound of 1.46. This means that, after adjustment for other confounders, an unmeasured or inadequately measured confounder would need to be associated with both step count and NAFLD risk by a hazard ratio of 1.52 to explain away the observed association, or a hazard ratio of 1.46 for the observed association confidence interval to include the null value. This represents a relatively large necessary association, as all included confounders had a hazard ratio below this threshold, excepting alcohol consumption.

## DISCUSSION

To our knowledge, this is the largest study of device-measured daily step count and risk of incident NAFLD, yielding three key findings. First, higher median daily step count was associated with lower risk of NAFLD, and this association is much stronger than has been previously reported.^4–12^ Second, this association was independent, but partially mediated by BMI, hypertension, medication use, and diabetes. Finally, the observed association was consistent, but attenuated, when considering MRI-based NAFLD assessments.

A striking result from the present analysis is a much stronger than previously reported steady decline in NAFLD risk with increasing daily step count in tandem with evidence of a log-linear dose-response relationship. While prior prospective studies have demonstrated this association, our study shows at least double the strength of the association. A meta-analysis of prior studies undertaken as part of this analysis demonstrated a fixed-effects pooled hazard ratio of 0.83 (0.81–0.85; I^2^: 93.12%; supplemental tables S3, S4, and S5 and supplemental figures S7 and S8) when comparing those who are most physically active to those who are least physically active. For comparison, our study demonstrated a hazard ratio of 0.35 (0.29–0.43), comparing those in the highest step count quarter to those in the lowest. This much stronger association may be explained by the use of objective physical activity measurements, as seven of the eight studies in the meta-analysis utilised questionnaire-based instruments to characterise activity.^4–8,10–12^ Indeed, a recent meta-analysis conducted by Ekeland and colleagues demonstrated that effect sizes for associations between physical activity and risk of death were twice as large for accelerometer-measured physical activity compared to self-reported physical activity.^25^ This probably reflects that objective instruments provide more valid, reliable, and comprehensive activity estimates.^14^ Indeed, in our analysis the observed activity-NAFLD effect size was halved when using a self-reported physical activity measurement compared to an accelerometer measurement.

Physical activity was independently associated with NAFLD risk; however, there were several relevant mediating factors, principally, obesity. One hypothesis for the observed association between physical activity and risk of NAFLD is that lower levels of activity lead to higher levels of adiposity, specifically in the form of visceral adipose tissue, leading to infiltration of fat into hepatocytes driven by free fatty acid efflux from adipose tissue.^6^ The present analysis demonstrated that adding BMI to the model attenuated the association between step count and risk of NAFLD, providing evidence to support this hypothesis. Other measures of central adiposity (body fat percentage, waist circumference) showed similar findings; given this consistency, we presented only findings using BMI due to its widespread use and ease of measurement. NAFLD is estimated to be present among 75-90% of individuals with overweight and obesity,^26^ therefore, modification of physical activity may confer less of a protective role among individuals with obesity.^2^ Diabetes, hypertension, and medication use may also mediate the association between activity and NAFLD. Prior studies have suggested a bidirectional association between insulin resistance and NAFLD^2^ and the DiRECT trial, involving a low-calorie diet intervention, recently demonstrated a link between hepatic fat loss and remission of diabetes.^27^ Mitra and colleagues postulate that NAFLD and diabetes are hallmarks of underlying metabolic dysfunction, and are the hepatic and pancreatic manifestations of a systemic disorder driven by insulin resistance and inflammation.^28^ Indeed, there is a recent discussion around refering to the combination of hepatic steatosis and metabolic dysfunction (type 2 diabetes, overweight, inflammation, hypertension) as metabolic-associated fatty liver disease (MAFLD) to reflect underlying disease processes.^29^

In a sensitivity analysis wherein NAFLD cases were ascertained using MRI-derived PDFF measures, the association between step count and incident NAFLD was attenuated in comparison to findings in the main analysis. One possible explanation for this attenuation is that NAFLD ascertainment using inpatient hospital records may identify more severe cases of NAFLD, while NAFLD ascertainment using PDFF measures may identify a wider spectrum of NAFLD presentation, including early, asymptomatic cases. Prevention of NAFLD progression is important, as fibrosis and cirrhosis, which are only present at more advanced stages, are the strongest predictors of adverse clinical outcomes, including cardiovascular disease complications and hepatocellular carcinoma.^30^ Importantly, estimates were comparable using either method among the cohort of participants with both accelerometer and imaging data; this consistency recommends the validity of our observed association.

Our study has several strengths including a large sample size, objective methods for measuring physical activity, two methods for ascertaining NAFLD, small numbers of participants lost to follow-up, and comprehensive collection and consideration of confounding variables and mediating factors. This analysis is limited by enrolment from only one geographic region with limited ethnic diversity, the possibility of inclusion of prevalent NAFLD cases, and evidence in the UK Biobank of a “healthy volunteer” bias, although previous studies in other areas suggest that many of the exposure-disease associations found in UK Biobank are largely generalisable to other populations.^31^ In addition, NAFLD ascertainment in our main analysis was based on record linkage with inpatient health records and the national death register, which may reflect an association between activity and ill health and may not capture all NAFLD cases. However, we performed an analysis accounting for possible ascertainment bias, and follow-up using MRI measurements showed comparable effect sizes to those using record linkage. Finally, although this analysis adjusted for relevant confounders as informed by a literature review, there may be residual confounding, although the calculated E-value for this analysis was moderately high at 1.52.

The present study demonstrates that accelerometer-measured physical activity is robustly associated with risk of NAFLD, and this association is approximately double the strength of that reported in studies using subjective activity measurements. Additionally, there is evidence that although this association is partially mediated by adiposity, physical activity does appear to have an independent role. These findings have important implications for reducing the global impact and burden of NAFLD, a disease affecting one third of the global population. More research is needed to replicate these findings in other settings; however, these results advocate the importance of activity for primary prevention of NAFLD and the need for clinical trials to endorse these findings.

## Contributors

ESF: conceptualisation, formal analysis, methodology, visualisation, writing - original draft; LP: conceptualisation, formal analysis, methodology, supervision, writing - review & editing; CH: conceptualisation, supervision, writing - review & editing; DP: conceptualisation, supervision, writing - review & editing; DB: conceptualisation, formal analysis, methodology, supervision, writing - review & editing; AD: conceptualisation, data curation, formal analysis, methodology, supervision, writing - review & editing.

## Data Sharing

The data used in this analysis is available to approved researchers from the UK Biobank.

## Data Availability

The data used in this analysis is available to approved researchers from the UK Biobank.

## Acknowledgements

We would like to thank the UK Biobank team and participants, without whom this research would not be possible.

## Funder Acknowledgements

For the purpose of open access, the author(s) has applied a Creative Commons Attribution (CC BY) licence to any Author Accepted Manuscript version arising.

## References

1. Riazi K, Azhari H, Charette JH, et al. The prevalence and incidence of NAFLD worldwide: a systematic review and meta-analysis. Lancet Gastroenterol Hepatol. Sep 2022;7(9):851–861. doi:10.1016/s2468-1253(22)00165-0

2. Byrne CD, Targher G. NAFLD: A multisystem disease. Journal of Hepatology. 2015/04/01/ 2015;62(1, Supplement):S47–S64. 10.1016/j.jhep.2014.12.012

3. Bugianesi E. Non-Alcoholic Fatty Liver Disease: A 360-degree Overview. 1 ed. Springer Cham; 2020:362.

4. Choi HI, Lee MY, Kim H, et al. Effect of physical activity on the development and the resolution of nonalcoholic fatty liver in relation to body mass index. BMC Public Health. Apr 5 2022;22(1):655. doi:10.1186/s12889-022-13128-6

5. Ge X, Wang X, Yan Y, et al. Behavioural activity pattern, genetic factors, and the risk of nonalcoholic fatty liver disease: A prospective study in the UK Biobank. Liver Int. Jun 2023;43(6):1287–1297. doi:10.1111/liv.15588

6. Kwak M-S, Kim D, Chung GE, Kim W, Kim JS. The preventive effect of sustained physical activity on incident nonalcoholic fatty liver disease. Liver International. 2017;37(6):919–926. 10.1111/liv.13332

7. Li P, Yang Q, Wang X, et al. Dose-response relationship between physical activity and nonalcoholic fatty liver disease: A prospective cohort study. Chin Med J (Engl). Jun 20 2023;136(12):1494–1496. doi:10.1097/cm9.0000000000002532

8. Lv Y, Ding D, Luo M, Tan X, Xia Y, Chen L. Physical Activity Intensity, Genetic Predisposition, and Risk of Non-alcoholic Fatty Liver Disease: A Prospective Cohort Study. Clin Gastroenterol Hepatol. Feb 2 2023;doi:10.1016/j.cgh.2023.01.011

9. Schneider CV, Zandvakili I, Thaiss CA, Schneider KM. Physical activity is associated with reduced risk of liver disease in the prospective UK Biobank cohort. JHEP Reports. 2021/06/01/ 2021;3(3):100263. 10.1016/j.jhepr.2021.100263

10. Sun S, Yang Q, Zhou Q, et al. Long-term exposure to air pollution, habitual physical activity and risk of non-alcoholic fatty liver disease: A prospective cohort study. Ecotoxicology and Environmental Safety. 2022/04/15/ 2022;235:113440. 10.1016/j.ecoenv.2022.113440

11. Sung K-C, Ryu S, Lee J-Y, Kim J-Y, Wild SH, Byrne CD. Effect of exercise on the development of new fatty liver and the resolution of existing fatty liver. Journal of Hepatology. 2016/10/01/ 2016;65(4):791-797. 10.1016/j.jhep.2016.05.026

12. Tsai Y-L, Chen SC-C. Joint effect of changes in physical activity and weight on incident non-alcoholic fatty liver disease. Journal of Epidemiology and Community Health. 2021;75(12):1215–1221. doi:10.1136/jech-2021-216728

13. Wang S-t, Zheng J, Peng H-w, et al. Physical activity intervention for non-diabetic patients with non-alcoholic fatty liver disease: a meta-analysis of randomized controlled trials. BMC Gastroenterology. 2020/03/12 2020;20(1):66. doi:10.1186/s12876-020-01204-3

14. Sallis JF, Saelens BE. Assessment of physical activity by self-report: status, limitations, and future directions. Res Q Exerc Sport. Jun 2000;71 Suppl 2:1–14. doi:10.1080/02701367.2000.11082780

15. Metabolic mediators of the effects of body-mass index, overweight, and obesity on coronary heart disease and stroke: a pooled analysis of 97 prospective cohorts with 1.8 million participants. The Lancet. 2014/03/15/ 2014;383(9921):970-983. 10.1016/S0140-6736(13)61836-X

16. Sudlow C, Gallacher J, Allen N, et al. UK Biobank: An Open Access Resource for Identifying the Causes of a Wide Range of Complex Diseases of Middle and Old Age. PLOS Medicine. 2015;12(3):e1001779. doi:10.1371/journal.pmed.1001779

17. Doherty A, Jackson D, Hammerla N, et al. Large Scale Population Assessment of Physical Activity Using Wrist Worn Accelerometers: The UK Biobank Study. PLOS ONE. 2017;12(2):e0169649. doi:10.1371/journal.pone.0169649

18. Small SR, Chan S, Walmsley R, et al. Development and Validation of a Machine Learning Wrist-worn Step Detection Algorithm with Deployment in the UK Biobank. medRxiv. 2023:2023.02.20.23285750. doi:10.1101/2023.02.20.23285750

19. Tudor-Locke C, Han H, Aguiar EJ, et al. How fast is fast enough? Walking cadence (steps/min) as a practical estimate of intensity in adults: a narrative review. British Journal of Sports Medicine. 2018;52(12):776–788. doi:10.1136/bjsports-2017-097628

20. Committee IR. Guidelines for data processing and analysis of the International Physical Activity Questionnaire (IPAQ)-short and long forms. http://www.ipaqkise/scoring.pdf. 2005;

21. Wilman HR, Kelly M, Garratt S, et al. Characterisation of liver fat in the UK Biobank cohort. PLOS ONE. 2017;12(2):e0172921. doi:10.1371/journal.pone.0172921

22. Easton DF, Peto J, Babiker AG. Floating absolute risk: an alternative to relative risk in survival and case-control analysis avoiding an arbitrary reference group. Stat Med. Jul 1991;10(7):1025–35. doi:10.1002/sim.4780100703

23. Rijnhart JJM, Lamp SJ, Valente MJ, MacKinnon DP, Twisk JWR, Heymans MW. Mediation analysis methods used in observational research: a scoping review and recommendations. BMC Medical Research Methodology. 2021/10/25 2021;21(1):226. doi:10.1186/s12874-021-01426-3

24. VanderWeele TJ, Ding P. Sensitivity Analysis in Observational Research: Introducing the E-Value. Annals of Internal Medicine. 2017/08/15 2017;167(4):268-274. doi:10.7326/M16-2607

25. Ekelund U, Tarp J, Steene-Johannessen J, et al. Dose-response associations between accelerometry measured physical activity and sedentary time and all cause mortality: systematic review and harmonised meta-analysis. BMJ. 2019;366:l4570. doi:10.1136/bmj.l4570

26. Cotter TG, Rinella M. Nonalcoholic Fatty Liver Disease 2020: The State of the Disease. Gastroenterology. 2020/05/01/ 2020;158(7):1851-1864. 10.1053/j.gastro.2020.01.052

27. Taylor R, Al-Mrabeh A, Zhyzhneuskaya S, et al. Remission of Human Type 2 Diabetes Requires Decrease in Liver and Pancreas Fat Content but Is Dependent upon Capacity for β Cell Recovery. Cell Metab. Oct 2 2018;28(4):547–556.e3. doi:10.1016/j.cmet.2018.07.003

28. Mitra S, De A, Chowdhury A. Epidemiology of non-alcoholic and alcoholic fatty liver diseases. Transl Gastroenterol Hepatol. 2020;5:16. doi:10.21037/tgh.2019.09.08

29. Gofton C, Upendran Y, Zheng MH, George J. MAFLD: How is it different from NAFLD? Clin Mol Hepatol. Feb 2023;29(Suppl):S17–s31. doi:10.3350/cmh.2022.0367

30. Ekstedt M, Hagström H, Nasr P, et al. Fibrosis stage is the strongest predictor for disease-specific mortality in NAFLD after up to 33 years of follow-up. Hepatology. May 2015;61(5):1547–54. doi:10.1002/hep.27368

31. Allen NE, Lacey B, Lawlor DA, et al. Prospective study design and data analysis in UK Biobank. Science Translational Medicine. 16(729):eadf4428. doi:10.1126/scitranslmed.adf4428

