## Supplementary Material for "Association of daily steps with incident non-alcoholic fatty liver disease: Evidence from the UK Biobank cohort"

### Table S1. ICD-10 and ICD-9 codes.

| **NAFLD** | |
| --- | --- |
| ICD-10 | |
| K75·8 | Other specified inflammatory liver diseases |
| K76·0 | Fatty (change of) liver, not elsewhere classified |
| ICD-9 | |
| 571·5 | Cirrhosis of liver without mention of alcohol |
| 571·8 | Other chronic nonalcoholic liver disease |
| 571·9 | Unspecified chronic liver disease without mention of alcohol |
| **Other Liver Disease** | |
| ICD-10 | |
| B16 | Acute hepatitis B |
| B16·0 | Acute hepatitis B with delta-agent (coinfection) with hepatic coma |
| B16·9 | Acute hepatitis B without delta-agent and without hepatic coma |
| B17 | Other acute viral hepatitis |
| B17·0 | Acute delta-(super) infection of hepatitis B carrier |
| B17·1 | Acute hepatitis C |
| B17·2 | Acute hepatitis E |
| B17·8 | Other specified acute viral hepatitis |
| B17·9 | Acute viral hepatitis, unspecified |
| B18 | Chronic viral hepatitis |
| B18·0 | Chronic viral hepatitis B with delta-agent |
| B18·1 | Chronic viral hepatitis B without delta-agent |
| B18·2 | Chronic viral hepatitis C |
| B18·8 | Other chronic viral hepatitis |
| B18·9 | Chronic viral hepatitis, unspecified |
| B19 | Unspecified viral hepatitis |
| B19·9 | Unspecified viral hepatitis without hepatic coma |
| C22·0 | Liver cell carcinoma |
| I82·0 | Budd Chiari syndrome |
| K70 | Alcoholic liver disease |
| K70·0 | Alcoholic fatty liver |
| K70·1 | Alcoholic hepatitis |
| K70·2 | Alcoholic fibrosis and sclerosis of liver |
| K70·3 | Alcoholic cirrhosis of liver |
| K70·4 | Alcoholic hepatic failure |
| K70·9 | Alcoholic liver disease, unspecified |
| K71 | Toxic liver disease |
| K71·0 | Toxic liver disease with cholestasis |
| K71·1 | Toxic liver disease with hepatic necrosis |
| K71·2 | Toxic liver disease with acute hepatitis |
| K71·3 | Toxic liver disease with chronic persistent hepatitis |
| K71·5 | Toxic liver disease with chronic active hepatitis |
| K71·6 | Toxic liver disease with hepatitis, not elsewhere classified |
| K71·7 | Toxic liver disease with fibrosis and cirrhosis of liver |
| K71·8 | Toxic liver disease with other disorders of liver |
| K71·9 | Toxic liver disease, unspecified |
| K73·2 | Chronic active hepatitis, not elsewhere classified |
| K73·9 | Chronic hepatitis, unspecified |
| K74·3 | Primary biliary cirrhosis |
| K74·4 | Secondary biliary cirrhosis |
| K74·5 | Biliary cirrhosis, unspecified |
| K75·4 | Autoimmune hepatitis |
| K76·5 | Hepatic veno-occlusive disease |
| K83·0 | Cholangitis |
| ICD-9 | |
| 070 | Viral hepatitis |
| 070·1 | Viral hepatitis A without mention of hepatic coma |
| 070·3 | Viral hepatitis B without mention of hepatic coma |
| 070·5 | Other specified viral hepatitis without mention of hepatic coma |
| 070·9 | Unspecified viral hepatitis without mention of hepatic coma |
| 155 | Malignant neoplasm of liver primary |
| 155·1 | Malignant neoplasm of intrahepatic bile ducts |
| 571·0 | Alcoholic fatty liver |
| 571·1 | Acute alcoholic hepatitis |
| 571·2 | Alcoholic cirrhosis of liver |
| 571·3 | Alcoholic liver damage, unspecified |
| 571·4 | Chronic hepatitis |
| 571·6 | Biliary cirrhosis |
| 277·62 | Other alpha-1-antitrypsin deficiency |
| 576·1 | Cholangitis |
| **Alcohol misuse** | |
| ICD-10 | |
| E24·4 | Alcohol-induced pseudo-Cushing's syndrome |
| F10·1 | Harmful use |
| F10·2 | Dependence syndrome |
| F10·3 | Withdrawal state |
| F10·4 | Withdrawal state with delirium |
| F10·5 | Psychotic disorder |
| F10·6 | Amnesic syndrome |
| F10·7 | Residual and late-onset psychotic disorder |
| F10·8 | Other mental and behavioural disorders |
| F10·9 | Unspecified mental and behavioural disorder |
| G31·2 | Degeneration of the nervous system due to alcohol |
| G62·1 | Alcoholic polyneuropathy |
| G72·1 | Alcoholic myopathy |
| I42·6 | Alcoholic cardiomyopathy |
| K29·2 | Alcoholic gastritis |
| T51·0 | Toxic effect of ethanol |
| T51·9 | Toxic effect of unspecified alcohol |
| X65·09 | Intentional self-poisoning by and exposure to alcohol, Home, During unspecified activity |
| Y57·3 | Adverse effects in therapeutic use: alcohol deterrents |
| Z50·2 | Alcohol rehabilitation |
| Z71·4 | Alcohol abuse counselling and surveillance |
| Z72·1 | Alcohol use |
| ICD-9 | |
| 291 | Alcoholic psychoses |
| 291·0 | Delerium tremens |
| 291·1 | Korsakov's psychosis alcoholic |
| 291·8 | Other specified alcoholic psychoses |
| 291·9 | Alcoholic psychoses unspecified |
| 303 | Alcohol dependence syndrome |
| 303·9 | Alcohol dependence syndrome |
| 305·0 | Nondependent alcohol abuse |
| 357·5 | Alcoholic polyneuropathy |
| 535·3 | Alcoholic gastritis |
| 980·0 | Toxic effect of ethyl alcohol |
| 980·9 | Toxic effect of unspecified alcohol |
| **Diabetes** | |
| ICD-10 | |
| E10 | Insulin-dependent diabetes mellitus |
| E10·0 | Insulin-dependent diabetes mellitus with coma |
| E10·1 | Insulin-dependent diabetes mellitus with ketoacidosis |
| E10·2 | Insulin-dependent diabetes mellitus with renal complications |
| E10·3 | Insulin-dependent diabetes mellitus with ophthalmic complications |
| E10·4 | Insulin-dependent diabetes mellitus with neurological complications |
| E10·5 | Insulin-dependent diabetes mellitus with peripheral circulatory complications |
| E10·6 | Insulin-dependent diabetes mellitus with other specified complications |
| E10·7 | Insulin-dependent diabetes mellitus with multiple complications |
| E10·8 | Insulin-dependent diabetes mellitus with unspecified complications |
| E10·9 | Insulin-dependent diabetes mellitus without complications |
| E11 | Non-insulin-dependent diabetes mellitus |
| E11·0 | Non-insulin-dependent diabetes mellitus with coma |
| E11·1 | Non-insulin-dependent diabetes mellitus with ketoacidosis |
| E11·2 | Non-insulin-dependent diabetes mellitus with renal complications |
| E11·3 | Non-insulin-dependent diabetes mellitus with ophthalmic complications |
| E11·4 | Non-insulin-dependent diabetes mellitus with neurological complications |
| E11·5 | Non-insulin-dependent diabetes mellitus with peripheral circulatory complications |
| E11·6 | Non-insulin-dependent diabetes mellitus with other specified complications |
| E11·7 | Non-insulin-dependent diabetes mellitus with multiple complications |
| E11·8 | Non-insulin-dependent diabetes mellitus with unspecified complications |
| E11·9 | Non-insulin-dependent diabetes mellitus without complications |
| E12 | Malnutrition-related diabetes mellitus with |
| E12·1 | Malnutrition-related diabetes mellitus with ketoacidosis |
| E12·3 | Malnutrition-related diabetes mellitus with ophthalmic complications |
| E12·5 | Malnutrition-related diabetes mellitus with peripheral circulatory complications |
| E12·8 | Malnutrition-related diabetes mellitus with unspecified complications |
| E12·9 | Malnutrition-related diabetes mellitus without complications |
| E13 | Other specified diabetes mellitus |
| E13·0 | Other specified diabetes mellitus with coma |
| E13·1 | Other specified diabetes mellitus with ketoacidosis |
| E13·2 | Other specified diabetes mellitus with renal complications |
| E13·3 | Other specified diabetes mellitus with ophthalmic complications |
| E13·4 | Other specified diabetes mellitus with neurological complications |
| E13·5 | Other specified diabetes mellitus with peripheral circulatory complications |
| E13·6 | Other specified diabetes mellitus with other specified complications |
| E13·7 | Other specified diabetes mellitus with multiple complications |
| E13·8 | Other specified diabetes mellitus with unspecified complications |
| E13·9 | Other specified diabetes mellitus without complications |
| E14 | Unspecified diabetes mellitus |
| E14·0 | Unspecified diabetes mellitus with coma |
| E14·1 | Unspecified diabetes mellitus with ketoacidosis |
| E14·2 | Unspecified diabetes mellitus with renal complications |
| E14·3 | Unspecified diabetes mellitus with ophthalmic complications |
| E14·4 | Unspecified diabetes mellitus with neurological complications |
| E14·5 | Unspecified diabetes mellitus with peripheral circulatory complications |
| E14·6 | Unspecified diabetes mellitus with other specified complications |
| E14·7 | Unspecified diabetes mellitus with multiple complications |
| E14·8 | Unspecified diabetes mellitus with unspecified complications |
| E14·9 | Unspecified diabetes mellitus without complications |
| ICD-9 | |
| 250 | Diabetes mellitus |
| 250·0 | Diabetes mellitus without mention of complication |
| 250·00 | Diabetes mellitus without mention of complication (adult-onset type) |
| 250·01 | Diabetes mellitus without mention of complication (juvenile type) |
| 250·09 | Diabetes mellitus without mention of compl. (adult/juvenile unspec.) |
| 250·1 | Diabetes with ketoacidosis |
| 250·10 | Diabetes with ketoacidosis (adult-onset type) |
| 250·11 | Diabetes with ketoacidosis (juvenile type) |
| 250·19 | Diabetes with ketoacidosis (adult/juvenile unspec.) |
| 250·2 | Diabetes with coma |
| 250·29 | Diabetes with coma (unspecified whether adult-onset or juvenile type) |
| 250·3 | Diabetes with renal manifestations |
| 250·4 | Diabetes with ophthalmic manifestations |
| 250·5 | Diabetes with neurological manifestations |
| 250·9 | Diabetes with unspecified complications |
| 250·99 | Diabetes with unspecified complications (unspecified onset) |
| **Hypertension** | |
| ICD-10 | |
| I10 | Essential (primary) hypertension |
| ICD-9 | |
| 401 | Essential hypertension |
| **Chronic lower respiratory disease** | |
| ICD-10 | |
| J41 | Simple and mucopurulent chronic bronchitis |
| J42 | Unspecified chronic bronchitis |
| J43 | Emphysema |
| J44 | Other chronic obstructive pulmonary disease |
| J45 | Asthma |
| J47 | Bronchiectasis |
| ICD-9 | |
| 490 | Bronchitis, not specified as acute or chronic |
| 491 | Chronic bronchitis |
| 492 | Emphysema |
| 493 | Asthma |
| 494 | Bronchiectasis |
| 496 | Chronic airways obstruction, not elsewhere classified |

ICD-10 and ICD-9 code selection informed by Hydes et al. ^44^ and Fairfield et al. ^45^

### Table S2. STROBE statement checklist for cohort studies.

|  | Item No | Recommendation | | Page No |
| --- | --- | --- | --- | --- |
| **Title and abstract** | 1 | (*a*) Indicate the study’s design with a commonly used term in the title or the abstract | | 1-2 |
|  |  | (*b*) Provide in the abstract an informative and balanced summary of what was done and what was found | |  |
| Introduction | | | | |
| Background/rationale | 2 | Explain the scientific background and rationale for the investigation being reported | | 4 |
| Objectives | 3 | State specific objectives, including any prespecified hypotheses | | 4-5 |
| Methods | | | | |
| Study design | 4 | Present key elements of study design early in the paper | | 5 |
| Setting | 5 | Describe the setting, locations, and relevant dates, including periods of recruitment, exposure, follow-up, and data collection | | 5-8 |
| Participants | 6 | (*a*) Give the eligibility criteria, and the sources and methods of selection of participants. Describe methods of follow-up | | 5-8; 10 |
|  |  | (*b*) For matched studies, give matching criteria and number of exposed and unexposed | |  |
| Variables | 7 | Clearly define all outcomes, exposures, predictors, potential confounders, and effect modifiers. Give diagnostic criteria, if applicable | | 5-8 |
| Data sources/ measurement | 8* | For each variable of interest, give sources of data and details of methods of assessment (measurement). Describe comparability of assessment methods if there is more than one group | | 5-8 |
| Bias | 9 | Describe any efforts to address potential sources of bias | | 5-11 |
| Study size | 10 | Explain how the study size was arrived at | | 5; 10 |
| Quantitative variables | 11 | Explain how quantitative variables were handled in the analyses. If applicable, describe which groupings were chosen and why | | 5-8 |
| Statistical methods | 12 | (*a*) Describe all statistical methods, including those used to control for confounding | | 5; 8-11 |
|  |  | (*b*) Describe any methods used to examine subgroups and interactions | |  |
|  |  | (*c*) Explain how missing data were addressed | |  |
|  |  | (*d*) If applicable, explain how loss to follow-up was addressed | |  |
|  |  | (*e*) Describe any sensitivity analyses | |  |
| Results | | |  | |
| Participants | 13* | (a) Report numbers of individuals at each stage of study—eg numbers potentially eligible, examined for eligibility, confirmed eligible, included in the study, completing follow-up, and analysed | | 11; Supplement p 13 |
|  |  | (b) Give reasons for non-participation at each stage | |  |
|  |  | (c) Consider use of a flow diagram | |  |
| Descriptive data | 14* | (a) Give characteristics of study participants (eg demographic, clinical, social) and information on exposures and potential confounders | | 11-15 |
|  |  | (b) Indicate number of participants with missing data for each variable of interest | |  |
|  |  | (c) Summarise follow-up time (eg, average and total amount) | |  |
| Outcome data | 15* | Report numbers of outcome events or summary measures over time | | 13-14 |
| Main results | 16 | (*a*) Give unadjusted estimates and, if applicable, confounder-adjusted estimates and their precision (eg, 95% confidence interval). Make clear which confounders were adjusted for and why they were included | | 13; supplemental p 14 |
|  |  | (*b*) Report category boundaries when continuous variables were categorized | | 12 |
|  |  | (*c*) If relevant, consider translating estimates of relative risk into absolute risk for a meaningful time period | | N/A (not appropriate) |
| Other analyses | 17 | Report other analyses done—eg analyses of subgroups and interactions, and sensitivity analyses | | 13-15 |
| **Discussion** |  |  | |  |
| Key results | 18 | Summarise key results with reference to study objectives | | 15 |
| Limitations | 19 | Discuss limitations of the study, taking into account sources of potential bias or imprecision. Discuss both direction and magnitude of any potential bias | | 18 |
| Interpretation | 20 | Give a cautious overall interpretation of results considering objectives, limitations, multiplicity of analyses, results from similar studies, and other relevant evidence | | 15-18 |
| Generalisability | 21 | Discuss the generalisability (external validity) of the study results | | 15-18 |
| **Other information** |  |  | |  |
| Funding | 22 | Give the source of funding and the role of the funders for the present study and, if applicable, for the original study on which the present article is based | | 20-21 |

Information on the STROBE Initiative is available at http://www.strobe-statement.org.

### Table S3. Literature review search terms.

|  | Element | Key word | Search terms in brief | EMBASE search terms | EMBASE Results | PubMed search terms | PubMed Results |
| --- | --- | --- | --- | --- | --- | --- | --- |
| 1 | **P**opulation | Adults | Search limited to Humans | Search limited to Humans |  |  |  |
| 2 | **E**xposure | Physical activity | Physical activity [Title] OR exercise [Title] OR walking [Title] OR  Steps per day [Title] OR step count [Title] OR movement [Title] | “physical activit*”.m_title OR exercise*.m_title OR walk*.m_titl OR “step count”.m_titl OR “steps per day”.m_titl OR movement.m_titl | 330,965 | ((((("physical activit*"[Title]) OR exercis*[Title]) OR walk*[Title]) OR "step count"[Title]) OR "steps per day"[Title]) OR "movement"[Title])) | 277,838 |
| 3 | **O**utcome | Incident NAFLD | NAFLD [Title] OR NASH [Title] OR MAFLD [Title] | NAFLD.m_titl OR NASH.m_titl OR MAFLD.m_titl OR “non-alcoholic fatty liver disease”.m_titl OR “nonalcoholic fatty liver disease” .m_titl OR “non-alcoholic steatohepatitis” .m_titl OR “nonalcholic steatohepatitis” .m_titl OR “metabolic-associated fatty liver disease” | 42,985 | (((((((NAFLD[Title]) OR NASH[Title]) OR MAFLD[Title]) OR "non-alcoholic fatty liver disease"[Title]) OR "nonalcoholic fatty liver disease"[Title]) OR "non-alcoholic steatohepatitis"[Title]) OR "nonalcoholic steatohepatitis"[Title]) OR "metabolic-associated fatty liver disease"[Title])) | 26,107 |
| 4 | **S**tudy Design | Prospective | Prospective OR Longitudinal OR Cohort OR Nested case-control OR Case cohort | Longitudinal study/ OR prospective study/ OR cohort analysis/ OR incidence/ OR attack rate/ OR cumulative incidence/ OR risk/ OR follow up/ OR “nested case-contol” .m_titl OR “cohort” .m_titl OR “case-cohort”.m_titl OR “longitudinal” .m_titl OR “prospective” .m_titl OR “follow-up”.m_titl | 4,185,643 | (((((((((((((("prospective studies"[MeSH Terms]) OR "longitudinal studies"[MeSH Terms]) OR "cohort studies"[MeSH Terms]) OR "incidence"[MeSH Terms]) OR "risk"[MeSH Terms]) OR "follow up studies"[MeSH Terms]) OR "nested case-control"[Title]) OR "cohort"[Title]) OR "case-cohort"[Title]) OR "longitudinal"[Title]) OR "prospective"[Title]) OR "follow-up"[Title])) | 3,759,817 |
| 5 |  |  | 2 AND 3 AND 4 |  | 77 |  | 127 |
| 6 |  |  | Limit (5) to (1) |  | 73 |  | 107 |

### Table S4. Summary of articles included in literature review.

| **Publication** | **Year Published** | **Sample Size** | **Country/Region** | **Exposure measure** | **Outcome measure** | **Relevant findings** | **Adjustments** |
| --- | --- | --- | --- | --- | --- | --- | --- |
| Choi | 2022 | N = 130,144  Body mass index (BMI) change > 0: N = 87,316  BMI change ≤ 0: N = 42,828 | South Korea | Korean version of the International Physical Activity Questionnaire Short Form (IPAQ-SF)  Mean age at assessment: 37·16 years | Ultrasound  Median duration of follow-up: 3·03 years | Most active vs. inactive (BMI change > 0) – HR: 0·94 (95% CI: 0·89 - 0·99)  Most active vs. inactive (BMI change ≤ 0) – HR: 0·88 (95% CI: 0·78 - 0·99) | Age, sex, centre, year of screening exam, smoking status, alcohol intake, education level, waist circumference, waist circumference changes |
| Ge | 2023 | N = 338,087 | United Kingdom | International Physical Activity Questionnaire  Mean age at assessment: 56.97 (8.06) years | Hospital inpatient records, death registration  Median duration of follow-up: 12.4 (11.7 – 13.1) years | Highest physical activity tertile vs. lowest physical activity tertile: 0.85 (95% CI: 0.77 – 0.93), P for trend <0·001 | Age, sex, smoking status, drinking status, education level, Townsend deprivation index, fruit and vegetable intake, red meat intake, BMI, metabolic health status, genetic risk categories, sedentary behaviour |
| Kwak | 2017 | N = 1,373 | South Korea | Korean version of the PA questionnaire from the National Health and Nutrition Examination Survey  Mean age at assessment: 51·4 ± 9·3 years | Ultrasound  Median duration of follow-up: 4·42 years | Fourth metabolic equivalent of task-min/week Quartile vs. First metabolic equivalent of task-min/week Quartile – HR: 0·66 (95% CI: 0·46 - 0·94), P for trend = 0·025 | Age, gender, body mass index, smoking, hypertension, diabetes, soft drink consumption, coffee consumption, waist circumference during follow-up, visceral adipose tissue, subcutaneous adipose tissue, homeostatic model assessment for insulin resistance |
| Li | 2023 | Fatty liver index (FLI) -defined NAFLD: N = 121,607  Hepatic steatosis index (HSI)-defined NAFLD: N = 118,946 | Taiwan | Self-reported questionnaire (not specified)  Mean age at assessment, FLI: 37·84 ± 11·91 years  Mean age at assessment, HSI: 38·39 ± 12·27 years | FLI and HSI  Median duration of follow-up, FLI: 3 (2 – 6) years  Median duration of follow-up, HSI: 3 (2 – 6) years | FLI: Very high physical activity vs. very low physical activity – HR: 0·71 (0·66 - 0·77), P for trend <0·001  HSI: Very high physical activity vs. very low physical activity – HR: 0·78 (95% CI: 0·73 - 0·84), P for trend <0·001 | Age, sex, education, body mass index, fruit intake, vegetable intake, fried food intake, season, smoking, alcohol consumption, strenuousness of work, occupational exposure, hypertension, diabetes, dyslipidaemia, cancer, and cardiovascular disease |
| Lv | 2023 | N = 288,495 | United Kingdom | International Physical Activity Questionnaire  Mean age at assessment: 56·2 ± 8·1 years | Hospital inpatient records, death registration, primary care data  Median duration of follow-up: 12·5 years | >50% vigorous-intensity physical activity vs. no vigorous-intensity physical activity – HR: 0·77 (95% CI: 0·68 - 0·86) | Age, sex, education, household income, Townsend deprivation index, assessment centres, smoking, healthy diet score, alcohol consumption, daily sedentary time, body mass index, total moderate to vigorous physical activity volume, baseline depression, dyslipidaemia, hypertension, diabetes, cardiovascular disease, cancer |
| Schneider | 2021 | N = 95,574 | United Kingdom | Wrist-worn triaxial accelerometer  Mean age at assessment: ~60 years (not specified for whole group) | Hospital inpatient records, death registration  Mean duration of follow-up: 5·5 years | Highest quartile of activity compared to lowest quartile – HR: 0·47 (95% CI: 0·28 - 0·77) | Body mass index, age, sex, alcohol consumption, month of accelerometry, smoking, hypertension, hyperlipidaemia, waist circumference, diabetes, overall health, sleep duration, wear-time, number of falls in previous year, mean kcal intake, mean sugar intake, mean fat intake, mean carbohydrate intake |
| Sun | 2022 | Fatty liver index (FLI)-defined NAFLD: N = 34,753 | Taiwan | Self-reported questionnaire (not specified)  Mean age at assessment, FLI: 40·4 ± 12·5 years  Mean age at assessment, HSI: 41·0 ± 12·9 years | FLI | Very high physical activity compared to very low physical activity – HR: 0·53 (95% CI: 0·48 - 0·58) | Age, year of enrolment, season of measurement, sex, smoking status, alcohol consumption, occupational exposure, educational attainment, vegetable intake, fruit intake, sugar drink, fried food intake, physical activity at work, cancer, long-term use of hyperlipidaemia drugs, cardiovascular disease, and hypertension |
| Sung | 2016 | N = 126,811 | South Korea | Korean version of the International Physical Activity Questionnaire Short Form (IPAQ-SF)  Mean age at assessment: 40·5 ± 9·7 years | Ultrasound  Mean duration of follow-up: 5 years | Exercise ≥5 times per week compared to no exercise – HR: 0·86 (95% CI: 0·8 - 0·92) | Age, sex, centre, year of screening exam, smoking status, alcohol intake, education level, body mass index, diabetes, hypertension, cardiovascular disease, change in body mass index |
| Tsai | 2021 | N = 47,058 | Taiwan | Self-reported questionnaire (not specified)  Mean age at assessment: ~33 years | Ultrasound  Mean duration of follow-up: 2·95 ± 1·13 years | Annual physical activity increase of 1 metabolic equivalent of task-hour/week compared to no change – HR: 0·88, (95% CI: 0·84 - 0·91) | Physical activity change, interaction between sex and physical activity change, sex, age, drinking, smoking, hypertension, fasting blood glucose, alanine aminotransferase, gamma-glutamyl transferase, triglycerides, total cholesterol, high-density lipoprotein cholesterol, body mass index, physical activity |

### Table S5. Literature review assessment of bias using the Newcastle-Ottawa Quality Assessment Scale for Cohort Studies.

| **Study** | **Representativeness of the exposed cohort** | **Selection of the unexposed cohort** | **Ascertainment of exposure** | **Demonstration that outcome of interest was not present at start of study** | **Comparability of cohorts on the basis of the design or analysis** | **Assessment of outcome** | **Was follow-up long enough for outcomes to occur (median/mean ≥ 3 years)** | **Adequacy of follow up cohorts** |
| --- | --- | --- | --- | --- | --- | --- | --- | --- |
| Choi et al., 2022 | Somewhat representative ★ | Drawn from same community as exposed ★ | Self-report questionnaire | Yes ★ | Study controls for age, sex, alcohol, and SES ★  Study controls for any additional factor ★ | Ultrasound, unclear if blinded and independent | Yes ★ | Yes ★ |
| Ge et al., 2022 | Somewhat representative ★ | Drawn from same community as exposed ★ | Self-report questionnaire | Yes ★ | Study controls for age, sex, alcohol, and SES ★  Study controls for any additional factor ★ | Record linkage ★ | Yes ★ | Yes ★ |
| Kwak et al., 2017 | Somewhat representative ★ | Drawn from same community as exposed ★ | Self-report questionnaire | Yes ★ | Study does not control for alcohol consumption  Study controls for any additional factor ★ | Ultrasound, unclear if blinded and independent | Yes ★ | Yes ★ |
| Li et al., 2023 | Somewhat representative ★ | Drawn from same community as exposed ★ | Self-report questionnaire | Yes ★ | Study controls for age, sex, alcohol, and SES ★  Study controls for any additional factor ★ | Blood-based test ★ | Yes ★ | Yes ★ |
| Lv et al., 2023 | Somewhat representative ★ | Drawn from same community as exposed ★ | Self-report questionnaire | Yes ★ | Study controls for age, sex, alcohol, and SES ★  Study controls for any additional factor ★ | Record linkage ★ | Yes ★ | Yes ★ |
| Schneider et al., 2021 | Somewhat representative ★ | Drawn from same community as exposed ★ | Accelerometer ★ | Yes ★ | Study does not control for SES  Study controls for any additional factor ★ | Record linkage ★ | Yes ★ | Yes ★ |
| Sun et al., 2022 | Somewhat representative ★ | Drawn from same community as exposed ★ | Self-report questionnaire | Yes ★ | Study controls for age, sex, alcohol, and SES ★  Study controls for any additional factor ★ | Blood-based test ★ | Yes ★ | Yes ★ |
| Sung et al., 2016 | Somewhat representative ★ | Drawn from same community as exposed ★ | Self-report questionnaire | Yes ★ | Study controls for age, sex, alcohol, and SES ★  Study controls for any additional factor ★ | Ultrasound, unclear if blinded and independent | Yes ★ | Yes ★ |
| Tsai et al., 2021 | Somewhat representative ★ | Drawn from same community as exposed ★ | Self-report questionnaire | Yes ★ | Study does not control for SES  Study controls for any additional factor ★ | Ultrasound, unclear if blinded and independent | No | Yes ★ |

### Figure S1. Exclusions to derive analysis population.

96,548 participants with quality accelerometer data

Poor wear time (n = 6,626)

Poor calibration (n = 111)

Very high overall activity (n = 68)

Very low overall activity (n = 6)

Prevalent liver disease based on hospital data (n = 737)

History of alcohol misuse based on hospital data (n = 1,770)

Missing covariate data (n = 3,006)

Missing lifestyle data (n = 1,536)

Missing sociodemographic data (n = 1,289)

Missing adiposity data (n = 181)

Loss to follow-up before accelerometer study entry (n = 4)

91,031 participants included in main analysis

103,359 participants with

accelerometer data

15,689 participants included in PDFF sensitivity analysis

Participants without imaging data (n = 75,123)

Participants with prevalent NAFLD based on PDFF* (n = 219)

*Participants had an imaging visit prior to accelerometer wear and had a PDFF>5.5%.

### Figure S2. Association of accelerometer-measured daily step count with risk of NAFLD, sequential adjustment of confounders.

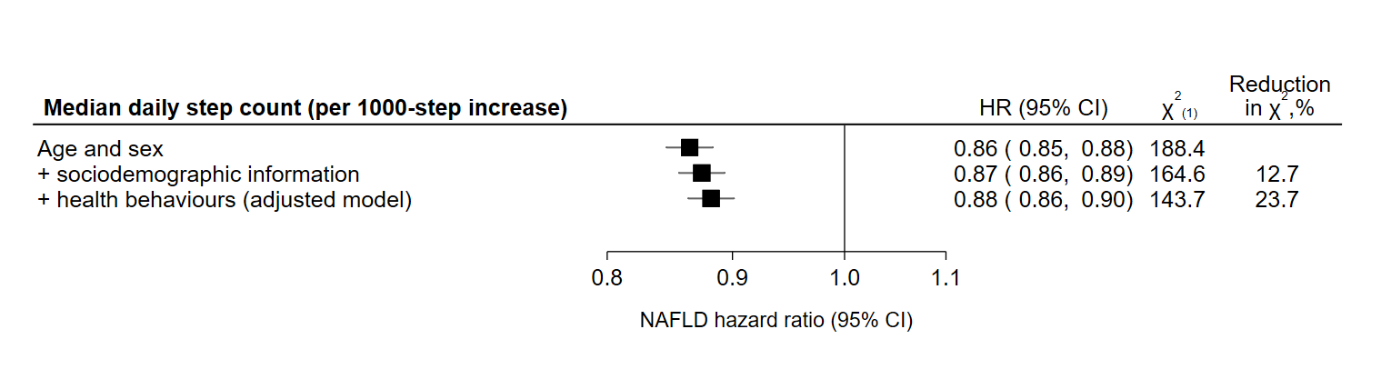

Sociodemographic variables: sex, ethnicity, Townsend deprivation index, educational attainment

Health behaviours: alcohol consumption, smoking status, fruit and vegetable consumption

Reduction in Χ^2^ is in relation to the minimally adjusted model (age and sex).

### Figure S3. Association of accelerometer-measured and self-reported physical activity with risk of NAFLD after a median of 7.9 years of follow-up in 76,464 UK Biobank participants.

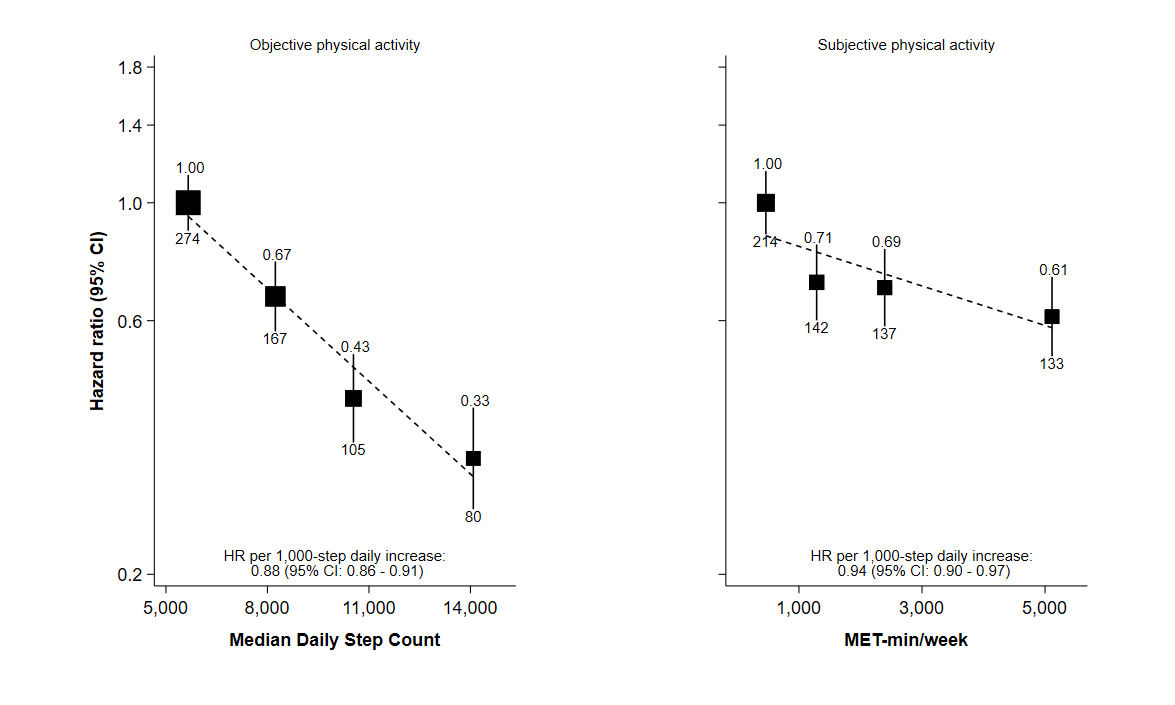

Adjusted for sex, ethnicity, Townsend deprivation index, educational attainment, alcohol consumption, smoking status, fruit and vegetable consumption and using age as the time scale; the number above each vertical line is the HR, and the number below each vertical line is the number of events; among participants with both subjective and objective activity measurements (N = 76,464).

### Figure S4. Association of quartiles of accelerometer-measured daily step count with risk of NAFLD, stratified by body mass index.

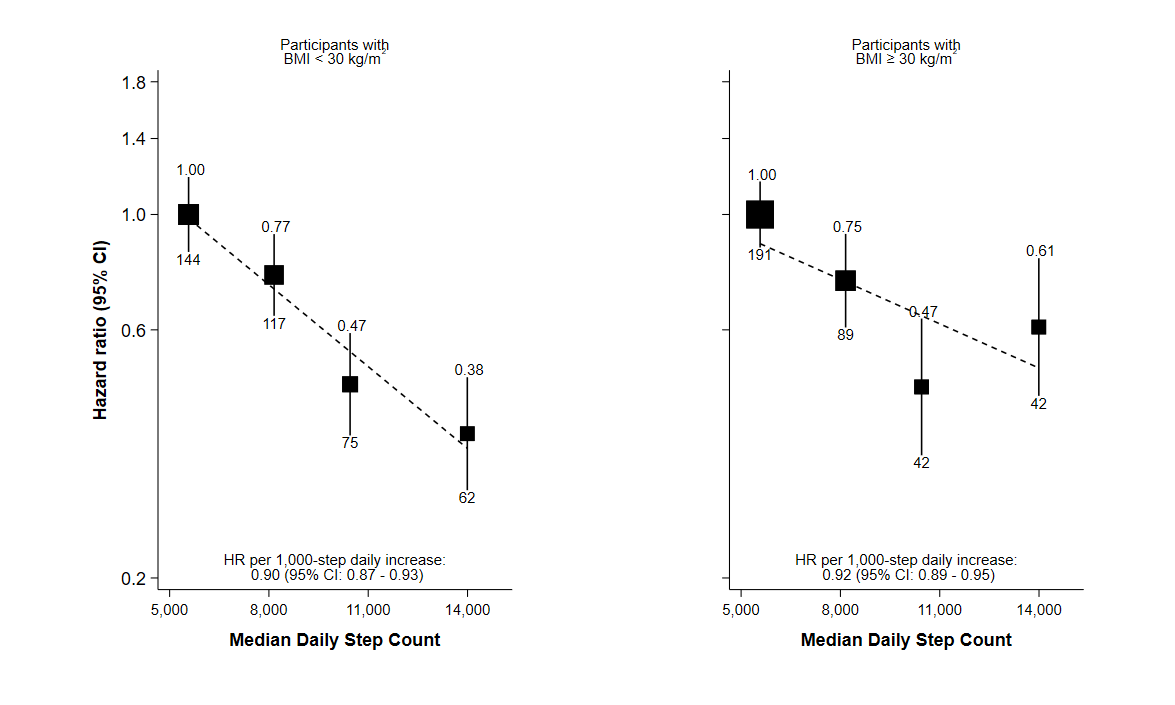

Adjusted for sex, ethnicity, Townsend deprivation index, educational attainment, alcohol consumption, smoking status, fruit and vegetable consumption and using age as the time scale; the number above each vertical line is the HR, and the number below each vertical line is the number of events.

### Figure S5. Association of quartiles of accelerometer-measured daily step count with risk of NAFLD, sensitivity analyses.

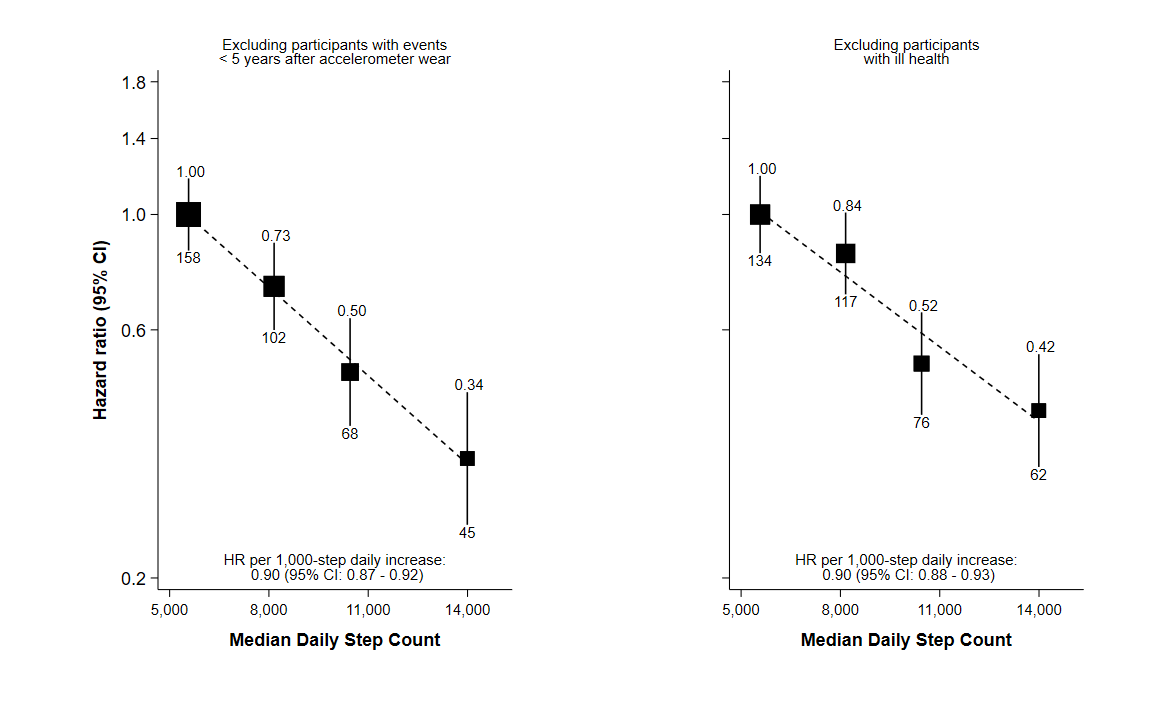

Adjusted for sex, ethnicity, Townsend deprivation index, educational attainment, alcohol consumption, smoking status, fruit and vegetable consumption and using age as the time scale; the number above each vertical line is the HR, and the number below each vertical line is the number of events.

Ill health is defined as either self-reported poor health or the presence of a co-morbid condition (cancer, chronic lower respiratory disease, diabetes, or hypertension). This analysis additionally adjusted for the number of hospital episodes in a participant’s record.

### Figure S6. Association of quartiles of accelerometer-measured daily step count with log(proton density fat fraction).

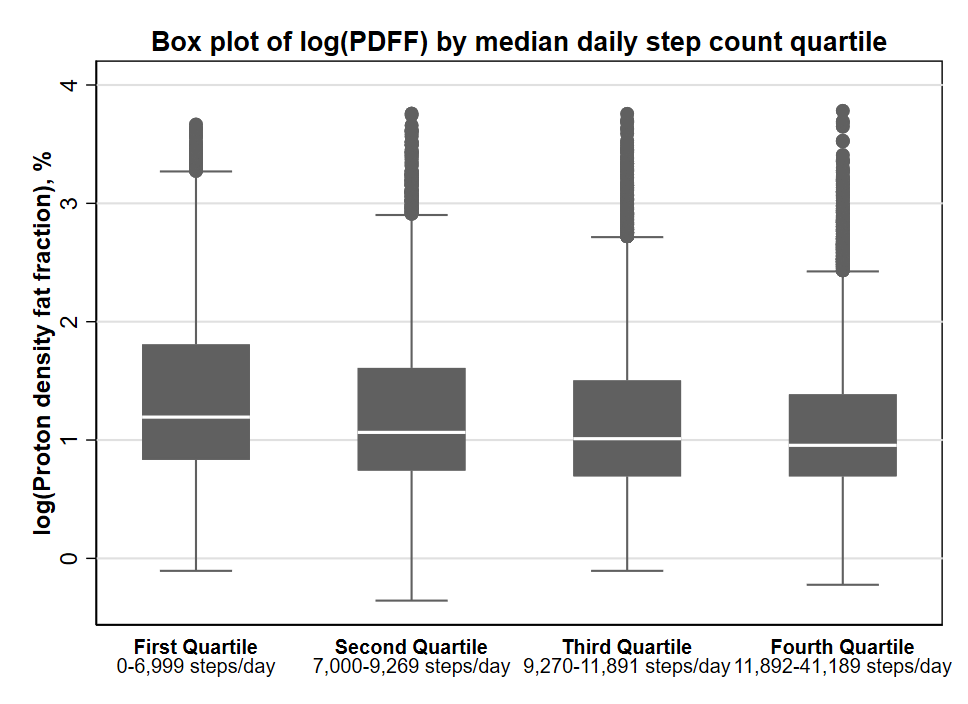

### Figure S7. PRISMA flow diagram of articles included in literature review.

Records screened and assessed for eligibility:

(n = 155)

Records removed *before screening*:

Duplicate records removed (n = 29)

Records excluded:

Study design not prospective (n = 74)

Prevalent NAFLD (n = 68)

NAFLD not main outcome (n = 3)

Paediatric population (n = 1)

Articles included in review:

(n = 9)

Records identified from:

EMBASE (n = 73)

PubMed (n = 107)

Citation tracking (4)

*From:* Page MJ, McKenzie JE, Bossuyt PM, Boutron I, Hoffmann TC, Mulrow CD, et al. The PRISMA 2020 statement: an updated guideline for reporting systematic reviews. BMJ 2021;372:n71. doi: 10·1136/bmj.n71

### Figure S8. Forest plot of prospective studies assessing the association between physical activity and risk of incident NAFLD, separated by method for assessing NAFLD.

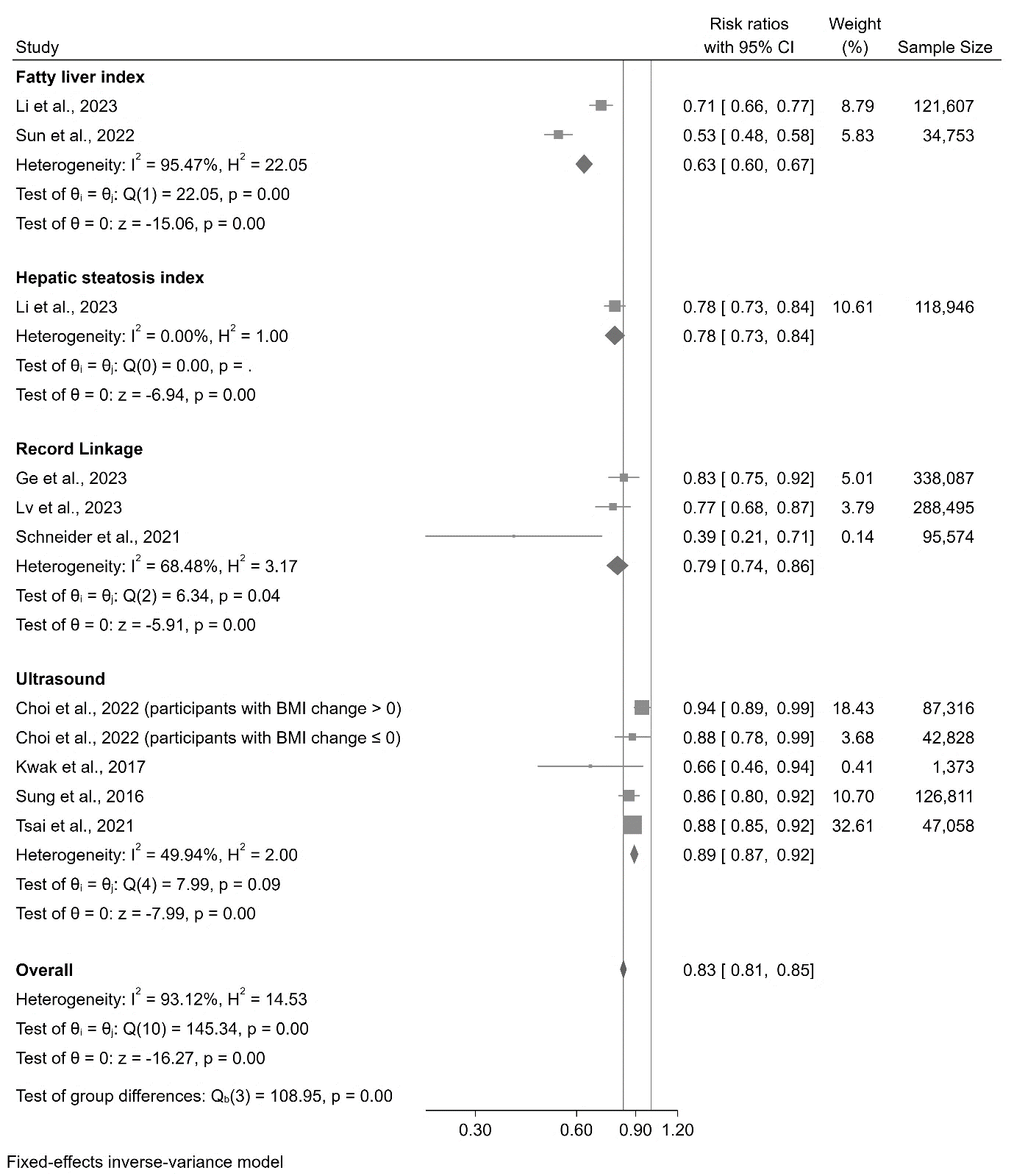

Highest level of activity compared to lowest level of activity.

All included studies used questionnaires to assess physical activity, except Schneider et al., which used a triaxial accelerometer.
